## Supplementary material for "Efficacy and safety of ethanolamine oleate in sclerotherapy in patients with difficult-to-resect venous malformations: A multicenter, open-label, single-arm study": Protocol

The protocol, informed consent form, case report form, and other materials related to this clinical trial (hereinafter referred to as "Clinical Trial Information") are confidential and will be provided only to those directly involved in this clinical trial (i.e., head of the institutes, coordinating office, investigators/subinvestigators, collaborators, drug administrators, and the Institutional Review Board). Except when explaining the contents of this clinical trial to subjects, this clinical trial-related information may not be disclosed to third parties or used for any purpose other than the purpose of this clinical trial, unless prior written consent is obtained from the person who intends to conduct the clinical trial on his/her own.

Summary of Protocol

| Title | A Multicenter, open-label, single-arm study to evaluate the efficacy and safety of sclerotherapy with monoethanolamine oleate for venous malformations |
| --- | --- |
| Clinical phase | III |
| Objectives | To evaluate the efficacy and safety of sclerotherapy in patients with difficult-to-resect venous malformations when treated with monoethanolamine oleate. |
| Trial design | Multicenter, open-label, single-arm |
| No. of patients | 44 cases (22 cystic lesions, 22 diffuse lesions) |
| Target disease | Difficult-to-resect venous malformations |
| Inclusion criteria | 1. No age limitation  2. Signed informed consent from patient or legal guardian(s) (In case age of patient is under 20 years old)  3. Venous malformations are considered difficult to remove, and sclerotherapy is considered first line treatment option by investigator/subinvestigator. Difficulty to remove means a high risk of functional dysfunction due to excision, or a loss of appearance to affect in daily life.  4. Patients who have one or more target venous malformations with a major axis of 30 mm or more in extremities, and 20 mm or more in head and neck region on MRI or CT  5. No thrombus and organized tissue that affects image evaluation or effect judgment in target venous malformations |
| Exclusion criteria | 1. Patients with multiple organ failure or DIC (Disseminated intravascular coagulation)  2. Patients who are taking or have taken drugs that may affect resolution of lesions (propranolol, herbal medicines [such as Kamishoyosan, Ninjinyoeito, Eppikajutsuto or Ogikenchuto], or sirolimus). However, patients can participate in the clinical study in case herbal medicines are discontinued 2 weeks or more before study drug administration.  3. Patients with diabetes mellitus with HbA1c>=8.0, or autoimmune disorder  4. Patients with liver dysfunction judged as grade C (10-15 points) in Child-Pugh classification  5. Patients with renal dysfunction with eGFR < 60mL/min/1.73m2  6. Patients with cardiac dysfunction with over grade II in NYHA classification  7. Patients having sclerotherapy within 6 months before obtaining informed consent  8. Patients with history of allergy to ethanolamine oleate or angiographic X-ray contrast agent as concomitant drugs  9. Patients having surgery over 45 minutes within 2 weeks before obtaining informed consent  10. Patients participating in other clinical study within 4 weeks before obtaining informed consent  11. Pregnant women, women who may be pregnant, or lactating women  12.Patients who are judged as inappropriate by investigator/subinvestigator |
| Test drug | Monoethanolamine oleate (investigational new ingredient symbol: FO-611) |
| Dose | Injection of 5% ethanolamine oleate which is double diluted by contrast or normal saline for the venous malformation, within maximum dosage of 0.4 mL/kg. Same method of administration is performed for children (under 15 years old). Maximum volume of the drug in once treatment is 30 mL after preparation. |
| Mode of administration | Intravenous administration (puncture directly into the lesion) |
| Duration of treatment | Single dose. However, for diffuse lesions, additional treatment (once) can be given as needed. |
| Observational period | Three months per case. However, in cases where additional treatment (once) was given, the period will be 3 months after the additional administration. |
| Efficacy endpoints | The efficacy evaluations below refer to the case of single administration. If additional treatment (one time) is given, evaluation is conducted after 3 months of additional administration (after 1 month of additional administration for 2) in "2.2 Other secondary endpoints" below) compared to that before the first administration.   1. Primary Endpoint   20% or more reduction in volume of venous malformations at the time of 3 months after intervention   2. Secondary endpoint   2.1 Significant secondary endpoint  Improvement in symptoms such as pain originated from venous malformations at the time of 3 months after intervention  2.2 Other secondary endpoints  1) Reduction rate in volume of venous malformations at the time of 3 months after intervention  2) Improvement in symptoms such as pain originated from venous malformations at the time of 1 months (4 weeks) after intervention  3) Improvement of QOL (ADL) at the time of 3 months after intervention   3. Other endpoint   Rate of change in appearance at the time of 3 months after intervention; much improvement, improvement, no change and worse. |
| Safety endpoints | Adverse events, side effects, laboratory values |
| Main statistical methods | Due to differences in disease characteristics, cystic and diffuse lesions will be analyzed separately as separate cohorts for all endpoints.   1. Primary endpoint:   For the number of subjects who achieve 20% or more reduction from baseline (before administration) in the volume of venous malformations at 3 months of treatment with the study drug, if the null hypothesis is "the proportion of the population achieving at least 20% reduction from baseline in the volume of venous malformations at 3 months of treatment with the study drug is less than 20%", a binomial distribution Tests based on the binomial distribution are performed. The significance level will be 2.5% one-sided. The 95% confidence interval between the proportion of subjects achieving at least 20% reduction from baseline in the volume of venous malformations at 3 months of treatment with the study drug and the proportion based on the score test will also be estimated.   2. Secondary endpoints:   2.1 Important secondary endpoints  For the symptom (pain) score associated with the target lesion at 3 months after sclerotherapy, estimate the median -1-fold change from baseline(before administration) to 3 months of sclerotherapy and its 95% confidence interval. Also, if the null hypothesis is "the median -1-fold change from baseline to 3 months post-sclerotherapy in the population is less than or equal to 0." and a Wilcoxon signed-rank test is performed. The significance level is set at 2.5% one-sided.  2.2 Other secondary endpoints  1) Estimate the median percent reduction from baseline (before administration) and its 95% confidence interval for the percent reduction (volume) of the target lesion at 3 months after sclerotherapy. Also, if the null hypothesis is "the median reduction in target lesion volume at 3 months after sclerotherapy is less than or equal to zero." The Wilcoxon signed-rank test is performed when the null hypothesis is "The median reduction rate (volume) of the target lesion at 3 months after sclerotherapy is less than 0. The significance level is set at 2.5% one-sided.  2) Estimate the median -1-fold change from baseline (before administration) to 1 month post sclerotherapy and its 95% confidence interval for the symptom (pain) score associated with the target lesion at 1 month (4 weeks) post sclerotherapy. Also, if the null hypothesis is "the median -1-fold change from baseline to one month after sclerotherapy in the population is less than or equal to zero." and a Wilcoxon signed-rank test is performed. The significance level is set at 2.5% one-sided.  3) Estimate the median change from baseline (before administration) to 3 months after sclerotherapy and its 95% confidence interval for the QOL score. We will also perform a Wilcoxon signed-rank test when the null hypothesis is "the median change from baseline to 3 months after sclerotherapy for the population is less than or equal to zero." and the Wilcoxon signed-rank test is performed when the null hypothesis is "The median change from baseline to 3 months after sclerotherapy in the population is less than or equal to zero. The significance level is set at 2.5% one-sided.  3. other evaluation items  Estimate the percentage of each category (obvious improvement, improvement, unchanged, and worsening) for the change in appearance at 3 months after sclerotherapy. We will also estimate the percentage of obvious improvement or betterment and its 95% confidence interval, with the null hypothesis being "the percentage of cases with obvious improvement or betterment is less than 20%." and then perform a test based on the binomial distribution. The significance level is set at 2.5% one-sided.  Main statistical methods  4. safety endpoints:  1) For each adverse event and adverse drug reaction, the number of subjects and incidence rate are shown. The number of subjects and the incidence rate by organ, symptoms and findings, and grade should be tabulated. For serious adverse events, the number of subjects and incidence rate should be indicated and listed.  2) Summarize summary statistics of laboratory measurements by time of survey. |
| Coordinating Committee | Tadashi Nomura, Department of Plastic and Reconstructive Surgery, Kobe University Hospital  Mine Ozaki, Department of Plastic and Reconstructive Surgery, Kyorin University Hospital |
| Institutions | 8 institutes:  Kyorin University Hospital, The University of Tokyo Hospital,  Juntendo University Urayasu Hospital, Keio University Hospital,  National Center for Child Health and Development,  Shinshu University Hospital, Osaka Medical and Pharmaceutical University Hospital, Kobe University Hospital |
| Trial period | Enrollment period: January 1, 2021 - March 31, 2023  Trial period: January 1, 2021 - June 30, 2023 |

Contents

Title page	1

[5.1	Efficacy endpoints](#_Toc117669595)	27

[5.1.1	Primary endpoint](#_Toc117669596)	27

[5.2.1.2	Collection and reporting of adverse events](#_Toc117669604)	31

[6.1.2	Diffuse lesions (if additional treatment is implemented)	3](#_Toc117669608)5

[6.2.4	Discontinuation](#_Toc117669620)	41

[6.2.4.1	Time of discontinuation](#_Toc117669621)	41

[6.2.4.2	3months after administration(12 weeks after administration) <discontinued cases> (Tolerance ±7 days)	42](#_Toc117669622)

[6.2.4.3	(In case of additional treatment) 3 months after additional administration (12 weeks after additional administration) <Discontinued cases	42](#_Toc117669623)

[8.6	Target number of cases and rationale for setting](#_Toc117669645)	47

Abbreviations (general)

| Abbreviation | Full spell |
| --- | --- |
| ADL | Activities of Daily Living |
| AMED | Japan Agency for Medical Research and Development |
| ARO | Academic Research Organization |
| CR | Complete Response |
| CRO | Contract Research Organization |
| CRF | Case Report Form |
| CSR | Clinical Study Report |
| CT | Computed Tomography |
| DIC | Disseminated Intravascular Coagulation |
| DSA | Digital Subtraction Angiography |
| eGFR | Estimated Glomerular Filtration Rate |
| EQ-5D | EuroQOL 5 dimensions |
| FAS | Full Analysis Set |
| GCP | Good Clinical Practice |
| IRB | Institutional Review Board |
| MRI | Magnetic Resonance Imaging |
| NYHA | New York Heart Association |
| PCA | Passive Cutaneous Anaphylaxis |
| PedsQL | Pediatric Quality of Life Inventory |
| pH | Potential of Hydrogen |
| PPS | Per Protocol Set |
| PR | Partial Response |
| QOL | Quality of Life |
| SAE | Serious Adverse Event |
| SAS | Safety Analysis Set |
| SpO_2_ | Saturaion of Percutaneous Oxygen |
| VAS | Visual Analogue Scale |

Abbreviations (Laboratories)

| Abbreviation | Full spell |
| --- | --- |
| Alb | Albumin |
| ALP | Alkaline Phosphatase |
| ALT | Alanine Aminotransferase |
| AMY | Amylase |
| APTT | Activated Partial Thromboplastin Time |
| AST | Aspartate Aminotransferase |
| BUN | Blood Urea Nitrogen |
| Ca | Calcium |
| CK | Creatine Kinase |
| Cl | Chlorine |
| Cre | Creatinine |
| CRP | C-Reactive Protein |
| FDP | Fibrinogen/Fibrin Degradation Products |
| Glu | Glucose |
| γ-GTP | gamma -Glutamyl Transpeptidase |
| HbA1c | Hemoglobin A1c |
| HDL-chol | High density lipoprotein- Cholesterol |
| IP | Inorganic Phosphorus |
| K | Potassium |
| LDH | Lactate Dehydrogenase |
| Na | Sodium |
| PT | Prothrombin Time |
| PT-INR | International Normalized Ratio of Prothrombin Time |
| T. Bil | Total Bilirubin |
| T-chol | Total Cholesterol |
| TG | Triglyceride |
| TP | Total Protein |
| UA | Uric Acid |

1. Introduction

1.1　Medical background

1) About Venous Malformations

Venous malformation is an abnormality in the formation of vascular vessels during the embryonic period and is associated with symptoms such as pain and movement disorders. The estimated number of patients in Japan (Ministry of Health, Labour and Welfare's Research and Study on Intractable Hemangioma, Hemangiopericytoma, Lymphangioma, Lymphangiomatosis and Related Diseases, Ministry of Health, Labour and Welfare's Research and Policy Project on Intractable Diseases in Fiscal 2014) is approximately 20,000. Half of the estimated 20,000 patients are considered to be difficult to resect, such as extensive cases or cases with muscle involvement^1)^, and sclerotherapy is the treatment of choice for these patients (number of applicable patients: approximately 10,000).

2) History of the development of this product for venous malformations

In Japan, monoethanolamine oleate, anhydrous ethanol, and polidocanol are used in off-label sclerotherapy. Regarding safety, although serious life-threatening side effects have been reported for anhydrous ethanol and polidocanol (Jo JY, et al: Cardiovascular collapse due to right heart failure following ethanol sclerotherapy, Frost et al: A case of a child with two cardiac arrests during polidocanol vascular sclerotherapy)^2, 3)^ , monoethanolamine oleate is considered a safe sclerosing agent with few side effects, with no serious side effects as shown in a systematic review by Horbach et al (venous malformation or lymphatic malformations). In 188 patients in 5 studies in diseases with venous malformations or lymphatic malformations, skin ulceration and skin necrosis [6 cases, 3%] were observed as side effects, but no systemic side effects, facial neuropathy, or other side effects were reported)^4)^. In a study that evaluated the cytotoxicity of each hardener on muscle tissue, ethanol was the most cytotoxic, with monoethanolamine oleate and polidocanol being comparable.^5)^ Efficacy was also reported with monoethanolamine oleate showing reduction of lesions and improvement of symptoms in 88-100% of cases^4)^, and in 83 other cases (85 operations) in which monoethanolamine oleate was administered mainly to children (average age 15.1 years: 3 months to 21 years of age) reported complete remission of symptoms in 79 lesions and significant improvement in 6 lesions.^6)^

Sclerotherapy is also a common treatment for venous malformations overseas, and the American textbook on vascular malformations, Vascular Anomalies, details sclerotherapy as one of the best treatments for venous malformations.^7)^ It also describes various sclerosing agents (including monoethanolamine oleate).^7)^ In addition, Sclerotherapy for venous malformation has been shown to be effective in the two guidelines such as “Guidelines and parameters: Percutaneous sclerotherapy for the treatment of head and neck venous and lymphatic malformations (VM and LM)” and “Guideline Diagnosis and Treatment of Venous Malformations (VMs)” in US, and in the guideline such as “the European guidelines for sclerotherapy in chronic venous disorders” in Europe, mainly Germany. Although there are a variety of sclerosing agents used, no sclerosing agent has been approved by authorities for venous malformations.

Based on the above, monoethanolamine oleate is considered the best agent as a curing agent in terms of efficacy and safety.

2. Rationale, purpose, and risk/benefit assessment of the clinical trial

2.1　Rationale for conducting clinical trials

The dose of monoethanolamine oleate administered in this study is within the previously approved range (gastric varices). In addition, no problematic events have been reported for venous malformations at that dosage and administration that differ from the known profile in the previously approved indication, and efficacy has already been reported in multiple literature reports, so we believe that additional nonclinical studies are unnecessary. On the other hand, many of the clinical studies reported to date have limited evidence on efficacy, with the endpoints being subjective. Therefore, we believe that it is necessary to conduct confirmatory clinical studies, including consideration of objective evaluation in assessing efficacy.

2.2　Purpose of the clinical trial

The purpose of this study is to evaluate the efficacy and safety of sclerotherapy in patients with difficult-to-resect venous malformations when treated with monoethanolamine oleate.

2.3　Risk/benefit

All subjects participating in this clinical trial are patients with difficult-to-resect venous malformations and will directly benefit from receiving a therapeutic agent for venous malformations, as there is currently no effective treatment for these malformations. In particular, improvement in quality of life (QOL), such as relief of pain associated with venous malformations, is expected.

All subjects participating in this clinical trial will receive the general medical benefit of careful and close observation by a health care professional during their participation in the clinical trial. Subject safety will be ensured through monitoring of adverse events based on clinical symptoms and laboratory tests. If the investigator or subinvestigator determines that there is a clinical concern, the safety of the subject will be given top priority.

In clinical studies of monoethanolamine oleate for venous malformations, tenderness and swelling at the injection site are known to be typical side effects, and Choi et al. reported local tenderness and swelling in all patients.^8)^ Other side effects included hemoglobinuria in 41.3-42.9%^9, 10)^, ulceration and skin necrosis in 3.0-14.7%^9-12)^, dysphagia in 7.7-12.5% of lesions near the buccal masseter muscle, and laryngeal edema in one case. Therefore, blood and urine tests should be performed the day after administration, and if hemoglobinuria or worsening of renal function is observed, these risks should be explained to the subject or surrogate, then further medical examination and treatment may be required.

Summary of Animal Experiments

Single-dose toxicity:

In rats, the fatal cases showed symptoms such as open mouth breathing, chain-stokes-like breathing, and clonic convulsions, and most of the deaths were due to respiratory paralysis. In dogs, urinary incontinence, tonic stretch spasms, and other symptoms were observed, and death occurred after respiratory distress.

Repeated dose toxicity :

A 4-week intravenous toxicity study in dogs was conducted at doses of 93, 31, 10, and 3 mg/kg every other day for 4 weeks (continuous administration was not possible due to site of administration swelling and hardening). No deaths occurred during the treatment period, but hematuria was observed in all groups, increased fibrinogen levels in the group of 31 mg/kg or higher, and decreased red blood cell counts, hematocrit and hemoglobin levels, increased white blood cell counts, and increased total bilirubin in the 93 mg/kg group. Pathological examination revealed thrombus formation and thickening of the vessel wall at the site of administration and granulosa cell infiltration of the dermis in the 3 mg/kg group and above, erythroblastic hyperplasia of the bone marrow and brown pigmentation of the renal tubular epithelium in the 31 mg/kg group and above, and thymic cortex atrophy and thrombus formation in the liver in the 93 mg/kg group. Based on the above, the toxicologically no-effect dose of the drug was estimated to be 10 mg/kg and the reliably toxic dose to be 93 mg/kg.

Antigenicity:

Results of systemic anaphylactic shock, PCA reaction test, and Schulzdale reaction test in guinea pigs were all negative.

Mutagenicity:

No mutagenicity was observed in reversion mutagenicity tests using bacteria (Salmonella, Escherichia coli) and mutagenicity tests using mammalian cultured cells by chromosome aberration tests.

Local disability:

　Local toxicity studies were performed in rabbits by intramuscular administration at doses of 5% and 0.05% concentration (1 mL). Gross findings showed mild white changes in all patients in the 0.05% group and moderate or greater white changes and mild to moderate hyperemia, hemorrhage, brownish changes, and swelling in all patients in the 5% group on day 2 after administration. On day 7 post-dose, there were no changes in the 0.05% group and mild to moderate white changes, swelling, and intense browning in all patients in the 5% group. Histological examination revealed dose-dependent edema of the hemorrhagic stroma, changes and necrosis of muscle fibers, and infiltration of histiocytes in both groups on post-dose day 2, as well as tumor and multinuclear cell infiltration in the 5% dose group. On day 7 after treatment, the above changes disappeared in the 0.05% group, and calcification of myofibers, fibroblast proliferation, and fibrosis were mildly observed. In the 5% treatment group, changes in hemorrhage myofibers, calcification, myoblast proliferation, fibrosis, moderate necrosis of myofibers, and fibroblast proliferation were observed.

3. Study design and study subjects population

3.1　Overall design and plan of the study

This is an open-label, multicenter, single-arm study to evaluate the efficacy and safety of monoethanolamine oleate (upper limit of 0.4 mL/kg as 5% monoethanolamine oleate) in difficult-to-resect venous malformations.

Subjects will be included in the study after written consent is obtained from the subject or surrogate. If screened and found eligible, the subject will be enrolled and administered monoethanolamine oleate. The observation period will be 3 months, and an efficacy test will be conducted after 3 months of treatment. If an additional treatment (one time) is administered, the efficacy evaluation will be conducted 3 months after the additional administration.

Termination of the study is defined as the date of last observation for the subject as a whole.

The overall clinical trial design is shown in Figure 3.1-1.

[Basis for setting up].

Because the difficult-to-resect venous malformations that are the subject of this study basically worsen without treatment, it is ethically difficult to establish a placebo group. Therefore, this clinical trial was an open-label, single-arm study in which only the actual drug group was administered. For the rationale for setting the target number of patients, see "8.6.2 Rationale for Setting the Target Number of Patients.

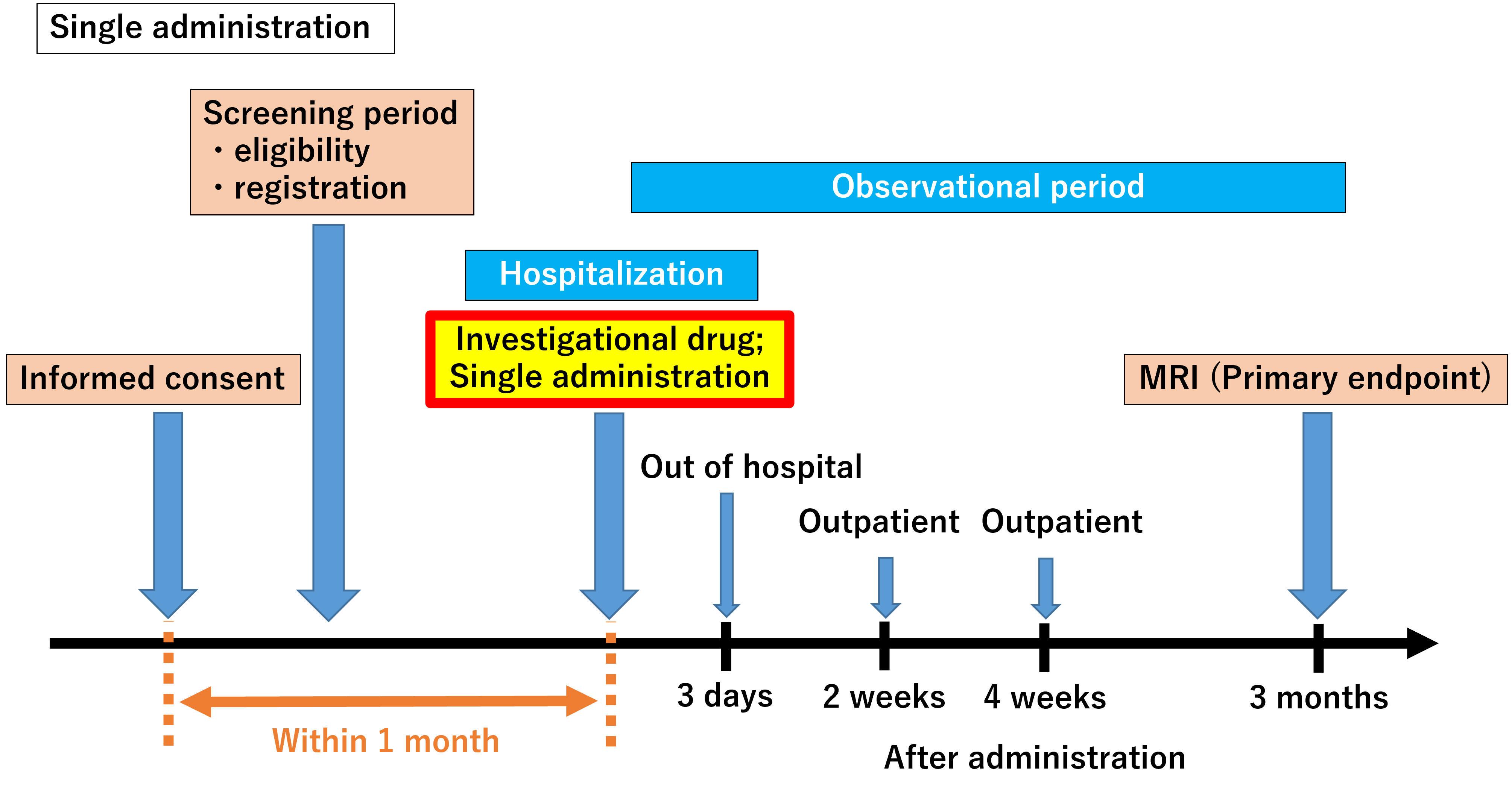

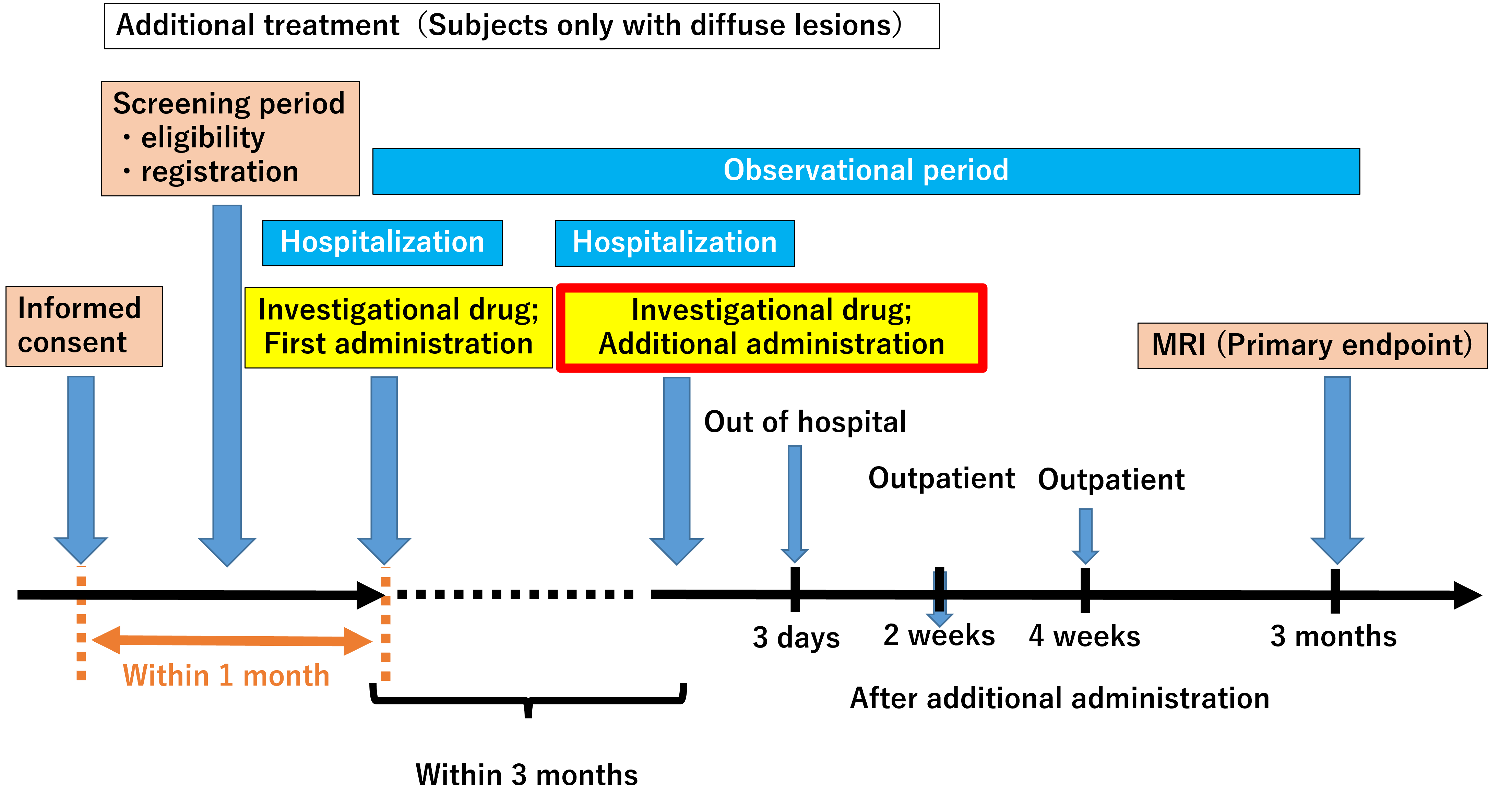

Figure 3. 1-1 Clinical trial design

3.2　Selection of the study population

3.2.1　Name of disease or diagnosis covered by this clinical trial

This clinical trial will be conducted in patients diagnosed with difficult-to-resect venous malformations.

3.2.2　Eligibility criteria

Patients who meet all of the following the inclusion criteria and do not violate any of the exclusion criteria will be eligible

3.2.2.1　Inclusion criteria

1. No age limitation
2. Signed informed consent from patient or legal guardian(s) (In case age of patient is under 20 years old)
3. Venous malformations are considered difficult to remove, and sclerotherapy is considered first line treatment option by investigator/subinvestigator. Difficulty to remove means a high risk of functional dysfunction due to excision, or a loss of appearance to affect in daily life.
4. Patients who have one or more target venous malformations with a major axis of 30 mm or more in extremities, and 20 mm or more in head and neck region on MRI or CT
5. No thrombus and organized tissue that affects image evaluation or effect judgment in target venous malformations

*: For patients 5 years of age or younger, the Clinical Trial Coordinating Committee will evaluate the appropriateness of inclusion or exclusion in advance.

3.2.2.2　Exclusion criteria

1. Patients with multiple organ failure or DIC (Disseminated intravascular coagulation)
2. Patients who are taking or have taken drugs that may affect resolution of lesions (propranolol, herbal medicines [such as Kamishoyosan, Ninjinyoeito, Eppikajutsuto or Ogikenchuto], or sirolimus). However, patients can participate in the clinical study in case herbal medicines are discontinued 2 weeks or more before study drug administration.
3. Patients with diabetes mellitus with HbA1c>=8.0, or autoimmune disorder
4. Patients with liver dysfunction judged as grade C (10-15 points) in Child-Pugh classification
5. Patients with renal dysfunction with eGFR < 60mL/min/1.73m2
6. Patients with cardiac dysfunction with over grade II in NYHA classification
7. Patients having sclerotherapy within 6 months before obtaining informed consent
8. Patients with history of allergy to ethanolamine oleate or angiographic X-ray contrast agent as concomitant drugs
9. Patients having surgery over 45 minutes within 2 weeks before obtaining informed consent
10. Patients participating in other clinical study within 4 weeks before obtaining informed consent
11. Pregnant women, women who may be pregnant, or lactating women
12. Patients who are judged as inappropriate by investigator/subinvestigator

3.2.2.3　Basis for establishing inclusion and exclusion criteria

Inclusion criteria

1: The target age for this clinical trial is not specified.

However, data collection for pain and QOL assessment will not be mandatory for patients aged 5 years or younger at the time of consent, as it is considered difficult to assess pain and QOL, which are part of the secondary endpoints. In the case of patients under 5 years of age, the Coordinating Committee will evaluate the appropriateness of inclusion or exclusion in advance.

4: In the field of plastic surgery, the size classification of the insurance procedure for tumor resection is 20 mm for the head and neck region and 30 mm for the extremities and trunk, and the same classification is used in actual clinical practice. In addition, the size at which detailed evaluation of venous malformation volume reduction or regression is possible on MRI is 20 mm or larger. Therefore, we judged it appropriate to set the target lesion size at 30 mm or larger in long diameter for the extremities and trunk and 20 mm or larger in long diameter for the head and neck.

2, 3: To select subjects who meet the objectives of the study, as well as to comply with GCP and ensure the safety of the subjects

5: Because it is difficult to evaluate images and determine effectiveness.

Exclusion criteria

1-7, 9-12: To take into account the safety of the subjects and to eliminate unexpected influences on the study results

8: To ensure the safety of the subject. However, when performing sclerotherapy for venous malformations, it is necessary to prepare the drug with an angiographic X-ray contrast medium. Therefore, if the patient is allergic to a contrast medium, it is necessary to select a contrast medium that is usable (does not cause allergy) for the patient, rather than making all contrast agents unusable. Since the subject is a patient who has no treatment options and whose ADL has declined due to pain and other symptoms, the above measures are acceptable from a risk-benefit perspective.

4. Treatment

4.1　Investigational drug

4.1.1　Source

The investigational drug will be provided by Fuji Chemical Industry Co., Ltd.

Please refer to "Appendix 1: 1.13 Investigational Drug Provider" for the person responsible for providing the investigational drug and contact information etc.

4.1.2　Feature

The characteristics of the investigational drug are as follows

Ingredient symbol: FO-611

Nonproprietary name: Monoethanolamine oleate

Dosage form: liquid/injection

Content: 1.000 g monoethanolamine oleate (0.822 g oleic acid, 0.178 g monoethanolamine) in 1 vial (10 g)

Description: Clear, viscous, colorless to pale yellow injectable liquid

Route of administration: intravenous (direct puncture into lesion)

4.1.3　Packaging and labeling of investigational drug

The investigational drug is contained in a vial and housed in an outer box. The outer box has a label indicating that the drug is investigational use, the identification code, protocol number, serial number, storage method, shelf life, and the name, affiliation, title, and address of the Coordinating Committee (chairman).

A sample of the labeling is shown in the figure below.

For Clinical Trials

FO-611

Protocol No.: EO-1

Serial number: P032A

Storage: Room temperature

Shelf life: 3 years

Coordinating Committee:

Kobe University Hospital, Department of Plastic and Reconstructive Surgery

Special Lecturer, Tadashi Nomura

7-5-2 Kusunoki-cho, Chuo-ku, Kobe, Hyogo 650-0017, Japan

4.1.4　Preparation methods and considerations

The following procedure is used to prepare the investigational drug.

1. Remove the flip-off cap of the vial of the investigational drug and clean the surface of the rubber stopper with alcohol cotton.
2. Take 10 mL of water for injection or angiographic X-ray contrast medium as a diluent in a syringe, and inject it into the vial by inserting a needle vertically into the center of the rubber stopper (20 mL volume after preparation).
3. After injection, shake the investigational drug horizontally to confirm that it is uniformly dissolved before use.

Suitability as a diluent is as follows

| Type of diluent | | Suitability as a diluent |
| --- | --- | --- |
| water for injection | | Suitable |
| physiological saline solution | | Unsuitable*. |
| X-ray contrast media for angiography | Iopamidol formulation (iodine content: 300, 370 mg/mL) | Suitable |
|  | Iohexol formulation (iodine content: 300, 350 mg/mL) | Suitable |
|  | I Oxaglic Acid Preparation | Unsuitable*. |
|  | Ioversol formulation (iodine content: 320, 350 mg/mL) | Unsuitable** |
|  | Iomeprol formulation (iodine content: 300, 350, 400 mg/mL) | Suitable |
|  | Iopromide preparation (iodine content: 300, 370 mg/mL) | Unsuitable** |

*: Do not use because it may become cloudy or not reduce viscosity.

**: Do not use as it may become cloudy.

The solution is a clear, viscous, colorless to pale yellow injectable solution.

The shaking during preparation causes minute air bubbles to be dispersed in the solution. After 1-3 minutes of standing, the air bubbles will collect on the surface of the solution, so aspirate the solution into a syringe, taking care to avoid air bubbles on the surface.

This product should be used immediately after mixing with water for injection or angiographic X-ray contrast medium. To avoid bacterial contamination, preparation should be done immediately before use, and residual liquid after use should not be reused.

4.1.5　Concentration of solution after preparation

Each vial (10 g: 10% monoethanolamine oleate) is dissolved in 10 mL of water for injection or angiographic X-ray contrast medium, and the solution concentration is 5% monoethanolamine oleate. The volume after preparation will be 20 mL due to double dilution.

4.1.6　Dosage and administration

A maximum dose of 0.4 mL/kg of 5% monoethanolamine oleate should be injected by puncture directly into the lesion. The same should be done for children (under 15 years of age). However, the maximum dose per treatment should be 30 mL (as prepared drug solution).

[Method of calculating the extreme volume (mL) of the dose]

(1) Measure the subject's body weight (kg) (to the first decimal place). If the second decimal place can be measured, it shall be rounded off and calculated to the first decimal place.

(2) Calculate the extreme volume (mL) of the dose using the following formula based on body weight. Round off the second decimal place and calculate to the first decimal place.

Extreme volume of dose (mL) = 0.4 (mL/kg) x body weight (kg)	<up to 30 mL>

4.1.7　Pre-treatment

Considering the use of contrast and sclerosing agents, 5-10 mL/kg of supplemental fluid should be administered prior to sclerotherapy.

4.1.8　Sclerotherapy procedure

Sclerotherapy is performed according to the following procedure

(1) In principle, general anesthesia using inhalation anesthesia should be administered to children (under 15 years of age). However, if the weight (physique) and pathological conditions of the patient are taken into consideration and it is judged that there is no problem, the same anesthesia method as that for adults may be used.

For adults (15 years of age and older), general anesthesia with anesthetics such as propofol and/or inhalation anesthesia should be used. It is also possible to perform the procedure under local anesthesia.

(2) The target lesion is confirmed using echocardiography or other means and punctured directly with a 22-25 G needle. Confirm that it is intravenous by checking for reverse bleeding.
(3) Then inject contrast medium and check the morphology and circulation of the lesion under DSA or fluoroscopy. Check for misinjection or problems on the outflow pathway.
(4) Once safety is confirmed, the sclerosing agent is injected.
(5) If there are areas within the target lesion where the sclerosing agent has not been injected sufficiently, repeat operations (2) to (4).
(6) The treatment is terminated when it is determined that the sclerosing agent has been injected into the target lesion.
(7) If the skin tone is checked and it is determined that there is a problem with skin circulation, a steroid-containing ointment should be applied.
(8) If necessary, use surgical tape or elastic bandages to compress and immobilize the treated area.
(9) The day after the sclerotherapy, release the compression and check the skin and lesion. Then, if necessary, reapply compression.

4.1.9　Post-treatment

For post-treatment, ointment (even steroid-containing ointment) or intravenous steroid infusion to reduce swelling in the treated area should be administered as needed. Surgical tape or elastic bandages should be applied as needed for approximately 2 weeks.

4.1.10　Additional treatment

Only for patients with diffuse lesions (when the target lesion is a diffuse lesion), if there is little improvement in symptoms such as pain after 4 weeks from the initial treatment (when there is almost no change in VAS [Visual Analogue Scale] or when an exacerbation is observed), and the investigator or subinvestigator determines that the additional treatment is needed, an additional administration (one time) will be performed (The need for additional treatment will be determined at least 4 weeks after the initial treatment). The dosage and administration of the additional treatment should be the same as that of the initial treatment. If additional treatment is administered, the efficacy and safety should be evaluated 3 months after the additional administration.

4.1.11　Duration of a drug administration

Single dose. However, if the target lesion is a diffuse lesion, additional administration (one time) may be given as needed.

4.1.12　Criteria for discontinuation of investigational drug

Discontinue the investigational drug if any of the following occur during administration of the investigational drug

1): Skin necrosis

If skin pallor is observed after the start of administration of the investigational drug, arterial embolization should be suspected and administration of the investigational drug should be discontinued immediately.

2): Immediate allergic reaction (anaphylaxis, etc.)

If an immediate allergic reaction related to the investigational drug is suspected, such as the appearance of a widespread rash or hemodynamic disruption after the start of administration of the investigational drug, administration of the investigational drug should be discontinued immediately.

4.1.13　Storage conditions for investigational drug

The investigational drug shall be stored in the packaging provided by the investigational drug provider and in accordance with the storage conditions indicated on the investigational drug label. Storage temperatures must be recorded.

Details of the conditions of the investigational drug management location and storage conditions of the investigational drug shall be separately stipulated in the "Procedures for Management of Investigational Drugs".

4.1.14　Investigational drug management

The investigational drug manager shall receive the investigational drug supplied by the investigational drug provider after ensuring that the following conditions are met

- The investigational review board (IRB) has approved the protocol for this clinical trial
- Notification to the regulatory authorities has been accepted.

The investigational drug manager at the site must record the status of investigational drugs received, inventory at the site, delivery to each subject, and return or disposal of unused investigational drugs. The date, quantity, serial number, expiry date, etc. must be entered in these records.

The investigational drug manager shall record that the investigational drug specified in the study protocol has been appropriately delivered to the subject, and shall check the consistency with the quantity of the investigational drug received from the investigational drug provider.

Details of the procedures for the management of investigational drugs are separately stipulated in the "Procedures for the Management of Investigational Drugs".

4.1.15　Precautions for delivery of investigational drug and at the time of delivery

The investigational drug manager delivers the investigational drug in accordance with the investigator's or subinvestigator's prescribing instructions.

4.1.16　Disposal of investigational drug

Since the investigational drug is essentially a single dose in this study, no collection of the investigational drug unused will occur. Any residual of the prepared investigational drug (unused residual solution) should be disposed of at each site. Since this investigational drug is regulated as a "deleterious substance," the investigator, subinvestigator or the investigational drug manager should properly dispose of the residual liquid in accordance with the disposal procedures for "deleterious substances" at each site.

If the prepared investigational drug is not used at all for some reason, it should be disposed of according to the same procedure. In this case, the investigational drug manager must record the reason why the drug was not used in the investigational drug management table.

4.2　Concomitant medications/adjunctive therapies, restrictions, emergency procedures

The investigator and subinvestigator will record details of concomitant medications and concomitant therapies from obtaining consent to the end of the observation period (until 3 months after the administration of the investigational drug, or until 3 months after the additional administration in the case of additional treatment) should be recorded in the case report form (CRF). The details to be recorded include the name of the concomitant drug, dosage, duration of administration, reason for use, etc. for the concomitant drug, and the name of the therapy, duration of therapy implementation, reason for implementation, etc. for the concomitant therapy.

4.2.1　Limitations on concomitant medications and concomitant therapies

In order to properly evaluate the effect of the investigational drug (monoethanolamine oleate), the concomitant use of polidocanol, propranolol, Chinese herbal medicines (Kamishoyosan, Ninjinyoeito, Eppikajutsuto, Ogikenchuto), and sirolimus is prohibited to be administered from the start date of administration of the investigational drug until the end of the observation period (until 3 months after administration of the investigational drug, or until 3 months after additional administration in the case of additional treatment).

4.2.2　Emergency medications, emergency procedures, additional concomitant medications/adjunctive therapies

If any of the adverse events listed in “4.1.12 Criteria for discontinuation of investigational drug” are observed during administration of monoethanolamine oleate, appropriate measures such as discontinuation of monoethanolamine oleate administration may be taken based on the judgment of the investigators and subinvestigators.

Any additional treatment (including blood transfusion, etc.) deemed necessary for the subject's health condition may be performed based on the judgment of the investigators and subinvestigators.

If hemolysis (gross hematuria) is observed, the patient should be treated with haptoglobin, if necessary, until the gross hematuria disappears. For adults (15 years of age and older), the initial dose is 2,000 units administered as a slow intravenous infusion. If symptoms do not improve, repeat doses may be administered as needed. The dosage may be adjusted according to the patient's age and body weight. The usual dosage for children (under 15 years of age) is 2,000 units per dose.

5.　Observation items and evaluation

5.1　Efficacy endpoints

The following efficacy evaluations are for single dosing. If an additional treatment (one time) is given, the results are evaluated 3 months after the additional dose (1 month after the additional dose for “5.1.2.2 Other secondary endpoint” #2), compared with the results before the first dose.

The calculation (evaluation) of the volume of venous malformations for the primary and secondary endpoints will be performed by the Central Judgment Committee independent from the Coordinating Committee, the investigator and the subinvestigator, based on the calculation method determined in advance by the Committee on Methods of Calculating Lesion Volume. Details of the Central Judgment Committee are separately stipulated in the "Procedures for the Central Judgment Committee.

5.1.1　Primary endpoint

Achievement of at least 20% reduction in volume of venous malformations from baseline (before administration) at 3 months of treatment with the investigational drug

[Basis for setting up].

Validity of the evaluation items:

In an ongoing Japanese investigator-initiated clinical trial of oral sirolimus treatment for lymphatic malformations, which is a similar disease of venous malformations, a 20% or greater reduction in the volume of the target lesion on MRI is considered a partial response (PR). Since the presence of a lesion can cause various symptoms, a reduction in volume of 20% or more is clinically significant. In addition, many literature studies have shown that patients with lesion reduction also experience improvement in pain.

In a trial conducted in the U.S. for oral sirolimus treatment of refractory vascular malformations, one of the endpoints was a 20% or greater reduction in the volume of the target lesion by MRI as PR, and the above domestic trial is based on this evaluation standard.

Validity of the evaluation period:

In evaluating the efficacy of sclerotherapy with monoethanolamine oleate, previous reports include Kaji et al.^5)^ evaluated by MRI at 3 months after treatment; Hoque et al.^6)^ evaluated at 8 weeks after sclerotherapy. A trial of gastric varices with a similar mechanism evaluated at 90 days after treatment. Based on the above, the evaluation period was set at 3 months.

Regarding safety, Nomura et al. reported^9)^ that skin necrosis or mucous membrane necrosis was observed in 14.7% of patients, but all of them improved with conservative treatment. The average time required for healing was 58.6 days (maximum about 90 days). Basically, the adverse drug reactions by sclerotherapy occur within a few days of treatment, and considering the period until the adverse drug reactions are cured as the necessary observation period, it is reasonable to set the evaluation period at 3 months.

5.1.2　Secondary endpoint(s)

5.1.2.1　Significant secondary endpoint

Improvement from baseline (before administration) in symptoms (pain) associated with the target lesion after 3 months of sclerotherapy*.

*: For pain rating scales, the VAS is used for patients aged 6 years and older; the Face scale is used for patients aged to 5 years, but the evaluation is not mandatory. For guardians, the VAS is used if the patient is 6 to 14 years old, and the Face scale is used if the patient is 5 years old or younger, and the evaluation should be done from the guardian’s perspective.. (The evaluation scale is shown in Appendix 2.)

For patients

| Patient Age | | 0 year old | 1 year old | 2-4 years old | 5 years old | 6-7 years old | 8-12 years old | 13-14 years old | 15 years and older |
| --- | --- | --- | --- | --- | --- | --- | --- | --- | --- |
| Pain | VAS |  |  |  |  | X | X | X | X |
|  | Face scale |  |  | X** | X** |  |  |  |  |

**: Evaluation is not mandatory.

For guardians

| Patient Age | | 0 year old | 1 year old | 2-4 years old | 5 years old | 6-7 years old | 8-12 years old | 13-14 years old | 15 years and older |
| --- | --- | --- | --- | --- | --- | --- | --- | --- | --- |
| Pain | VAS |  |  |  |  | X | X | X |  |
|  | Face scale | X | X | X | X |  |  |  |  |

[Basis for setting up].

In diffuse lesions, we considered it particularly important to show improvement in pain, so we set this as a significant secondary endpoint.

5.1.2.2　Other secondary endpoints

The efficacy endpoints are as follows:

1) Reduction rate in volume of venous malformations at the time of 3 months after intervention

2) Improvement in symptoms such as pain originated from venous malformations at the time of 1 month (4 weeks) after intervention^＊^

3) Improvement of QOL (ADL) at the time of 3 months after intervention^＊＊^

*: For pain rating scales, the VAS is used for patients aged 6 years and older; the Face scale is used for patients aged 2 to 5 years, but the evaluation is not mandatory. For guardians, the VAS is used if the patient is 6 to 14 years old, and the Face scale is used if the patient is 5 years old or younger to evaluate from the guardian’s perspective. (The evaluation scale is shown in Appendix 2.)

**: For the QOL evaluation scales, the EQ-5D Japanese version is used for patients aged 15 years and older, and the PedsQL Japanese version for patients aged 5 to 14 years, but the evaluation is not mandatory for patients aged 5 years. For guardians, the PedsQL Japanese version (proxy evaluation) is used if the patient is 14 years old or younger, and the evaluation should be done from the guardian’s perspective. The selection of the QOL evaluation scale was determined based on the review of the QOL Evaluation Method Review Committee. (The evaluation scale is shown in Appendix 2.)

For patients

| Patient Age | | 0 year old | 1 year old | 2-4 years old | 5 years old | 6-7 years old | 8-12 years old | 13-14 years old | 15 years and older |
| --- | --- | --- | --- | --- | --- | --- | --- | --- | --- |
| Pain | VAS |  |  |  |  | X | X | X | X |
|  | Face scale |  |  | X***. | X***. |  |  |  |  |
| QOL | EQ-5D |  |  |  |  |  |  |  | X |
|  | PedsQL |  |  |  | X***. | X | X | X |  |

****: Evaluation is not mandatory.

For guardians

| Patient Age | | 0 year old | 1 year old | 2-4 years old | 5 years old | 6-7 years old | 8-12 years old | 13-14 years old | 15 years and older |
| --- | --- | --- | --- | --- | --- | --- | --- | --- | --- |
| Pain | VAS |  |  |  |  | X | X | X |  |
|  | Face scale | X | X | X | X |  |  |  |  |
| QOL | PedsQL | X | X | X | X | X | X | X |  |

5.1.3　Other endpoint

Rate of change in appearance at the time of 3 months after intervention; much improvement, improvement, no change and worse.

5.2　Safety endpoints

1) Adverse events and Adverse drug reactions
2) Clinical laboratory test

5.2.1　Adverse event assessment

5.2.1.1　Definition of adverse events

Adverse event

An adverse event is defined as any unfavorable medical events that occur in patients from the time of informed consent is obtained until the end of the observation period (up to 3 months after the administration of the investigational drug, or up to 3 months after the administration of an additional dose in the case of additional treatment), regardless of whether or not it is causally related to the treatment.

Serious adverse events

Serious adverse events (SAEs) are defined as follows:

- Things that lead to death
- Life Threatening
- Requires hospitalization or extended hospitalization for treatment
- Permanent or marked disability or dysfunction
- Those that result in congenital anomalies
- Adverse events that are considered serious for other reasons include medically significant events that, based on appropriate medical judgment, may endanger the subject or require medical or surgical procedures that do not meet the above definition.

The term "life-threatening" in this definition means that the subject was at risk of death when the event occurred, not in the hypothetical sense that death might have resulted had the event been more severe.

Newly developed cancer must be reported as a serious adverse event regardless of the time between discontinuation of the investigational drug and the development of the cancer.

Adverse events that may lead to disability should be reported as serious adverse events as other medically significant conditions.

Notable Significant Adverse Events

The following events are defined as significant adverse events of interest for the safety profile.

Skin necrosis occurring after administration

Visual impairment occurring after administration (in case of facial lesions)

If a significant adverse event occurs, the investigator or subinvestigator shall promptly inform the Coordinating Committee via E-mail. Conduct the clinical trial with caution, referring to "4.1.12 Criteria for discontinuation of investigational drug" and "4.2.2 Emergency medications, emergency procedures, additional concomitant medications/adjunctive therapies"

Severity of adverse events

The severity of the adverse event will be determined by the following criteria and recorded in the CRF

Mild: easily tolerated signs or symptoms

Moderate: Signs or symptoms that interfere with daily life

Severe: Signs or symptoms of severe impairment to the extent that daily living is no longer possible.

Causal relationship with investigational drug

The causal relationship between the investigational drug and the adverse event shall be determined and recorded in the CRF using the following criteria, taking into consideration all relevant factors including the pattern of occurrence of the adverse event, temporal relationship, circumstances when the investigational drug was discontinued or re-administered, and confounding factors such as concomitant therapy, complications, and history of the event.

Causal relationship exists: a reasonable causal relationship exists between the administered investigational drug and the adverse event.

No causal relationship: other than above

5.2.1.2　Collection and reporting of adverse events

Collection of adverse events

The investigator and subinvestigator will collect and record the following adverse events on the CRF

- From the time of informed consent is obtained until the end of the observation period for each subject (up to 3 months after the administration of the study drug. In the case of additional treatment, until 3 months after the additional administration):

All adverse events (serious and non-serious)

- After the completion of the individual subject's clinical trial:

When investigators and subinvestigators become aware of SAEs that cannot be ruled out as causally related to the investigational drug, they must report these events promptly. The period for collecting information shall be until the completion of this entire clinical trial (until the last observation date of the last subject).

Reporting of adverse events to the head of the site, the Coordinating Committee, and the investigational drug provider, and the procedures for reporting adverse events

(1) When an SAE occurs, the investigator and subinvestigator must immediately report it to the head of the site and to the Coordinating Committee using the unified forms "Form 12 (Report on Serious Adverse Events)" and "Form for Detailed Description".
(2) The Coordinating Committee shall review the content of the SAE obtained from the investigator or subinvestigator and notify the investigators at other sites of the information regarding the SAE.
(3) The investigator at each site shall review the content of the SAE information obtained from the Coordinating Committee, discuss it with the Coordinating Committee as necessary, and report his/her opinion as the investigator (including the necessity of reporting to the MHLW) to the Coordinating Committee.
(4) The Coordinating Committee shall report promptly to the investigational drug provider by e-mail using the same form as in (1).
(5) When additional information on these events is obtained, a SAE reporting form should be submitted as an additional report. The same rules and procedures will be followed for the additional information as for the initial report of information.

Reporting of adverse events to the MHLW and its procedures

The Coordinating Committee must report to the MHLW within the established time limit if the case falls under a reportable case (SAE for which a causal relationship with the investigational drug cannot be denied) as stipulated in Article 273 of the Enforcement Regulations of the Pharmaceutical Affairs Law.

Required Information

For each adverse event, the investigator and subinvestigator shall record the necessary information (onset date, disappearance date, severity, treatment, outcome, severity, treatment with the investigational drug, etc.) in the CRF. In addition, investigators and subinvestigators must determine the causal relationship between all adverse events and the investigational drug.

The following events will also be treated as adverse events

・Aggravation of primary or pre-existing disease

・Fluctuations in vital signs, electrocardiograms, physical findings, and laboratory values that the investigator and subinvestigators consider clinically problematic

If such abnormalities existed prior to inclusion in the clinical trial, they are considered the baseline condition of the subject.

All adverse events, including those that persist at the end of a clinical trial for an individual subject, should be followed to the extent possible until the subject recovers to the state before the adverse event, or until the event is fully investigated or no new information is available.

6.　Investigational plan

6.1　Observation/testing items and schedule

Table 6.1.1-1 and Table 6.1.2-1 show the observation/test items and their implementation schedule for this clinical trial. The investigators and subinvestigators will conduct observations and inspections according to the schedule.

6.1.1　Cystic and diffuse lesions (No additional treatment is needed)

Table 6.1.1-1. Observation/examination items and implementation schedule

|  | Before treatment | | Treatment and observation period | | | | | | Discontinuation | |
| --- | --- | --- | --- | --- | --- | --- | --- | --- | --- | --- |
|  | Before administration  (Screening) | 1 day  before administration | Date of administration  (single dose) | 1 day  after administration | 3 days  after administration | 2 weeks  after administration | 4 weeks  after administration | 3months after administration  (12 weeks after administration) | Time of discontinuance | 3months after administration  (12 weeks after administration) |
| Tolerance (days) |  |  |  |  | ±1 | ±5 | ±5 | ±7 |  | ±7 |
| medical care category | outpatient | hospitalization | hospitalization | hospitalization | hospitalization | outpatient | outpatient | outpatient | hospitalization/  outpatient^1)^ | outpatient |
| Obtaining Consent | X ^2)^ |  |  |  |  |  |  |  |  |  |
| Eligibility assessment | X ^3)^ |  |  |  |  |  |  |  |  |  |
| Registration | X |  |  |  |  |  |  |  |  |  |
| Vital signs^4)^ |  | X |  | X | X | X | X | X | X | X |
| Body weight (usu. one's own) |  | X |  |  |  |  |  |  |  |  |
| Appearance^5)^ | X |  |  |  |  |  |  | X | X | X |
| Administration of an investigational drug |  |  | X ^6)^ |  |  |  |  |  |  |  |
| Clinical Laboratory (Blood)^7)^ | X ^8)^ |  |  | X ^9)^ |  |  |  |  | X |  |
| Urinalysis ^10)^ | X ^8)^ |  |  | X ^9)^ |  |  |  |  | X |  |
| Subjective symptoms (pain)^11)^ | X ^12)^ | X ^12)^ |  | X | X | X | X ^13)^ | X ^12)^ | X | X ^12)^ |
| MRI ^14)^ | X ^8)^ |  |  |  |  |  |  | X ^15)^ |  | X ^15)^ |
| QOL (including ADL)^16)^ | X |  |  |  |  |  | X | X | X | X |
| Eye check-up |  |  |  | X ^17)^ |  |  |  |  |  |  |
| Adverse event | X | X | X | X | X | X | X | X | X | X |
| Concomitant medications | X | X | X | X | X | X | X | X | X | X |

MRI: magnetic resonance imaging, QOL: quality of life, ADL: activities of daily living

1) Inpatient or outpatient, depending on the circumstances at the time of discontinuation.

2) Consent should be obtained within 1 month (30 days) prior to administration of the investigational drug.

3) After obtaining consent, screening tests are conducted to determine eligibility. If the subject is found to be eligible, the subject will be enrolled.

4) Blood pressure, pulse rate, body temperature, SpO_2_

5) Photograph the lesion site when observing the appearance.

6) Based on the patient's body weight on the day before dosing, a maximum dose of 0.4 mL/kg of 5% monoethanolamine oleate should be administered as the extreme dose (maximum is 30 mL).

7) The volume of blood collected is approximately 11 mL (maximum volume). The laboratory tests to be performed are red blood cell count, hemoglobin, hematocrit, white blood cell count, platelet count, white blood cell fraction (neutrophils, lymphocytes, eosinophils, basophils, monocytes), PT, APTT, PT-INR, fibrinogen, antithrombin III, FDP, D-dimer, Na, K, Cl, Ca, IP, BUN, Cre, UA, AST, ALT, LDH, γ-GTP, T.Bil, TP, Alb, ALP, CK, TG, T-chol, HDL-chol, AMY, CRP, Glu

8) Any laboratory data (within 30 days prior to obtaining consent) performed in the clinical setting unrelated to this study may be adopted as pre-dose data. However, for MRI, as a general rule, this shall be the case when imaging conditions specified in the "Imaging Evaluation Procedures" are used. In addition, if consent is obtained again for some reason, it is acceptable to use the MRI data as preadministration data if the initial MRI images were taken within 90 days from the date of consent.

9) If clinically relevant abnormalities are observed in clinical laboratory tests (blood) and urinalysis, such tests should also be performed 3 days after administration.

10) Urinalysis items (qualitative) are sugar, protein, urobilinogen, occult blood, leukocytes, pH, and specific gravity.

11) For pain rating scales, the VAS is used for patients 6 years of age and older; the Face scale is used for patients 2 to 5 years of age, but the evaluation is not mandatory. For guardians, the VAS is used if the patient is 6 to 14 years old, and the Face scale is used if the patient is 5 years old or younger to evaluate from the guardian’s perspective. (The evaluation scale is shown in Appendix 2.)

For patients

| Patient Age | | 0 year old | 1 year old | 2-4 years old | 5 years old | 6-7 years old | 8-12 years old | 13-14 years old | 15 years and older |
| --- | --- | --- | --- | --- | --- | --- | --- | --- | --- |
| pain | VAS |  |  |  |  | X | X | X | X |
|  | Face scale |  |  | X* | X* |  |  |  |  |

*: Evaluation is not mandatory.

For guardians

| Patient Age | | 0 year old | 1 year old | 2-4 years old | 5 years old | 6-7 years old | 8-12 years old | 13-14 years old | 15 years and older |
| --- | --- | --- | --- | --- | --- | --- | --- | --- | --- |
| pain | VAS |  |  |  |  | X | X | X |  |
|  | Face scale | X | X | X | X |  |  |  |  |

12) At "pre-dose (screening)", "1 day before administration", and "3 months after administration (12 weeks after administration)", two assessments of pain are performed: "last 24 hours" and "last 1 week" (other visits are only "last 24 hours").

13) In patients with diffuse lesions, if there is lack of improvement in pain after 4 weeks from the initial treatment and the investigator or subinvestigator determines that it is therapeutically necessary, an additional treatment (one time) may be administered within 3 months (the need for additional treatment will be determined at least 4 weeks after the initial treatment (The need for additional treatment will be determined at least 4 weeks after the initial treatment). The dosage and administration of the additional treatment should be the same as that of the initial treatment.

14) Additional CT scan or ultrasound may be performed as needed.

15) For MRI imaging only, the acceptable range after 3 months of administration is ±14 days.

16) For the QOL evaluation scales, the EQ-5D Japanese version is used for patients aged 15 years and older, and the PedsQL Japanese version for patients aged 5 to 14 years. But the evaluation is not mandatory for patients aged 5 years. For guardians, the PedsQL Japanese version (proxy evaluation) should be used when the patient is 14 years old or younger, and the evaluation should be done from the guardian’s perspective. (The evaluation scale is shown in Appendix 2.)

For patients

| Patient Age | | 0 year old | 1 year old | 2-4 years old | 5 years old | 6-7 years old | 8-12 years old | 13-14 years old | 15 years and older |
| --- | --- | --- | --- | --- | --- | --- | --- | --- | --- |
| QOL | EQ-5D |  |  |  |  |  |  |  | X |
|  | PedsQL |  |  |  | X** | X | X | X |  |

**: Evaluation is not mandatory.

For guardians

| Patient Age | | 0 year old | 1 year old | 2-4 years old | 5 years old | 6-7 years old | 8-12 years old | 13-14 years old | 15 years and older |
| --- | --- | --- | --- | --- | --- | --- | --- | --- | --- |
| QOL | PedsQL | X | X | X | X | X | X | X |  |

17) If periocular lesions are targeted, an ophthalmologic examination and examination should be performed the day after sclerotherapy.

6.1.2　Diffuse lesions (if additional treatment is implemented)

In diffuse lesions, an additional treatment (one time) may be administered if the initial dose is insufficient to alleviate pain (when there is almost no change in VAS or when an exacerbation is observed) and the investigator or subinvestigator determines that an additional dose is necessary (the need for additional treatment will be determined at least 4 weeks after the initial treatment).

During the period between the initial administration and the implementation of additional treatment, observation and examination should be conducted according to the schedule in the previous section "6.1.1 Cystic and diffuse lesions (No additional treatment is needed)".

Table 6.1.2-1. Observation/examination items and implementation schedule

|  | Before treatment | | Treatment and observation period | | | | | | | | Discontinuation | |
| --- | --- | --- | --- | --- | --- | --- | --- | --- | --- | --- | --- | --- |
|  | Before administration  (Screening) | 1 day  before administration | Date of administration (first dose) | ^5)^  ... | Date of administration (additional administration) | 1 day after additional administration | 3 days after additional administration | 2 weeks after additional administration | 4 weeks after additional administration | 3 months after additional administration  (12 weeks after) | Time of discontinuance | 3 months after additional administration  (12 weeks after) |
| Tolerance (days) |  |  |  |  |  |  | ±1 | ±5 | ±5 | ±7 |  | ±7 |
| medical care category | outpatient | hospitalization | hospitalization |  | hospitalization | hospitalization | hospitalization | outpatient | outpatient | outpatient | hospitalization/  outpatient^1)^ | outpatient |
| Obtaining Consent | X ^2)^ |  |  |  |  |  |  |  |  |  |  |  |
| Eligibility assessment | X ^3)^ |  |  |  |  |  |  |  |  |  |  |  |
| Registration | X |  |  |  |  |  |  |  |  |  |  |  |
| Vital signs^4)^ |  | X |  |  |  | X | X | X | X | X | X | X |
| Body weight (usu. one's own) |  | X |  |  | X ^6)^ |  |  |  |  |  |  |  |
| Appearance^7)^ | X |  |  |  |  |  |  |  |  | X | X | X |
| Administration of an investigational drug |  |  | X ^8)^ |  | X ^8)^ |  |  |  |  |  |  |  |
| Clinical Laboratory (Blood)^9)^ | X ^10)^ |  |  |  |  | X ^11)^ |  |  |  |  | X |  |
| Urinalysis^12)^ | X ^10)^ |  |  |  |  | X ^11)^ |  |  |  |  | X |  |
| Subjective symptoms (pain)^13)^ | X ^14)^ | X ^14)^ |  |  |  | X | X | X | X | X ^14)^ | X | X ^14)^ |
| MRI ^15)^ | X ^10)^ |  |  |  |  |  |  |  |  | X ^16)^ |  | X ^16)^ |
| QOL (including ADL)^17)^ | X |  |  |  |  |  |  |  | X | X | X | X |
| Eye check-up |  |  |  |  |  | X ^18)^ |  |  |  |  |  |  |
| Adverse event | X | X | X |  | X | X | X | X | X | X | X | X |
| Concomitant medications | X | X | X |  | X | X | X | X | X | X | X | X |

MRI: magnetic resonance imaging, QOL: quality of life, ADL: activities of daily living

1) Inpatient or outpatient, depending on the circumstances at the time of discontinuation.

2) Consent should be obtained within 1 month (30 days) prior to administration of the investigational drug.

3) After obtaining consent, screening tests are conducted to determine eligibility. If the subject is found to be eligible, the subject will be enrolled.

4) Blood pressure, pulse rate, body temperature, SpO_2_

5) During the period between the first dose and the implementation of additional treatment, observation and examination should be conducted according to the schedule in the previous section "6.1.1 Cystic and diffuse lesions (No additional treatment is not needed)".

6) In the case of an additional dose, the patient's weight should be measured on the day before the dose.

7) Photograph the lesion site when observing the appearance.

8) Based on body weight on the day before dosing, administer 0.4 mL/kg of 5% monoethanolamine oleate as the extreme dose
 (maximum is 30 mL).

9) The volume of blood collected is approximately 11 mL (maximum volume). The laboratory tests to be performed are red blood cell count, hemoglobin, hematocrit, white blood cell count, platelet count, white blood cell fraction (neutrophils, lymphocytes, eosinophils, basophils, monocytes), PT, APTT, PT-INR, fibrinogen, antithrombin III, FDP, D-dimer, Na, K, Cl, Ca, IP, BUN, Cre, UA, AST, ALT, LDH, γ-GTP, T.Bil, TP, Alb, ALP, CK, TG, T-chol, HDL-chol, AMY, CRP, Glu

10) Laboratory data (within 30 days prior to obtaining consent) performed in the clinical setting unrelated to this study are acceptable for adoption as pre-dose data. However, for MRI, as a general rule, this shall be the case when imaging conditions specified in the "Imaging Evaluation Procedures" are used. In addition, if consent is obtained again for some reason, it is acceptable to use the MRI data as preadministration data if the initial MRI images were taken within 90 days from the date of consent.

11) If clinically relevant abnormalities are observed in clinical laboratory tests (blood) and urinalysis, such tests should also be performed 3 days after administration.

12) Urinalysis items (qualitative) are sugar, protein, urobilinogen, occult blood, leukocytes, pH, and specific gravity.

13) For pain rating scales, the VAS is used for patients 6 years of age and older; the Face scale is used for patients 2 to 5 years of age, but the evaluation is not mandatory. For guardians, the VAS is used if the patient is 6 to 14 years old, and the Face scale is used if the patient is 5 years old or younger to evaluate from the guardian’s perspective. (The evaluation scale is shown in Appendix 2.)

For patients

| Patient Age | | 0 year old | 1 year old | 2-4 years old | 5 years old | 6-7 years old | 8-12 years old | 13-14 years old | 15 years and older |
| --- | --- | --- | --- | --- | --- | --- | --- | --- | --- |
| pain | VAS |  |  |  |  | X | X | X | X |
|  | Face scale |  |  | X* | X* |  |  |  |  |

*: Evaluation is not mandatory.

For guardians

| Patient Age | | 0 year old | 1 year old | 2-4 years old | 5 years old | 6-7 years old | 8-12 years old | 13-14 years old | 15 years and older |
| --- | --- | --- | --- | --- | --- | --- | --- | --- | --- |
| pain | VAS |  |  |  |  | X | X | X |  |
|  | Face scale | X | X | X | X |  |  |  |  |

14) At "pre-dose (screening)", "1 day before administration", and "3 months after additional administration (12 weeks after additional administration)", two assessments of pain are performed: "last 24 hours" and "last 1 week" (the other VISIT is "last 24 hours" only).

15) Additional CT scan or ultrasound may be performed if necessary.

16) For MRI imaging only, the allowable range after 3 months of additional administration is ±14 days.

17) For the QOL assessment scale, the EQ-5D Japanese version should be used for patients aged 15 years and older, and the PedsQL Japanese version should be used for patients aged 5 to 14 years. However, the evaluation is not mandatory for patients aged 5 years. For guardians, the PedsQL Japanese version (proxy evaluation) should be used when the patient is 14 years old or younger, and the evaluation should be done from the guardian’s perspective. (The evaluation scale is shown in Appendix 2.)

For patients

| Patient Age | | 0 year old | 1 year old | 2-4 years old | 5 years old | 6-7 years old | 8-12 years old | 13-14 years old | 15 years and older |
| --- | --- | --- | --- | --- | --- | --- | --- | --- | --- |
| QOL | EQ-5D |  |  |  |  |  |  |  | X |
|  | PedsQL |  |  |  | X** | X | X | X |  |

**: Evaluation is not mandatory.

For guardians

| Patient Age | | 0 year old | 1 year old | 2-4 years old | 5 years old | 6-7 years old | 8-12 years old | 13-14 years old | 15 years and older |
| --- | --- | --- | --- | --- | --- | --- | --- | --- | --- |
| QOL | PedsQL | X | X | X | X | X | X | X |  |

18) If periocular lesions are targeted, an ophthalmologic examination and examination should be performed the day after sclerotherapy.

6.2　Details of procedures for each implementation period

Detailed procedures for each implementation period are described below.

For the assessment of pain, the VAS is used for patients 6 years of age and older; the Face scale is used for patients 2 to 5 years of age, but the assessment is not mandatory. For guardians, the VAS is used if the patient is 6 to 14 years old, and the Face scale is used if the patient is 5 years old or younger to evaluate from the guardian’s perspective. EQ-5D Japanese version for patients aged 15 years and older, and the PedsQL Japanese version for patients aged 5 to 14 years (assessment is not mandatory for patients aged 5 years). For guardians, the PedsQL Japanese version (proxy evaluation) is used to evaluate the QOL from the guardian’s perspective if the patient is 14 years old or younger.

6.2.1　Before administration (screening)

After obtaining consent from the subject or surrogate, the subject's eligibility for participation in this clinical trial will be determined through screening observations and examinations in accordance with "3.2.2 Eligibility criteria".

With regard to "clinical examination," "urinalysis," and "MRI (in principle, when imaging is performed under the imaging conditions of the Imaging Evaluation Procedures)," if there are observation and examination results obtained within one month (30 days) prior to obtaining consent, they may be adopted as screening data after obtaining consent from the subject or surrogate. In addition, if consent is obtained again for some reason, it is acceptable to adopt the MRI images initially taken as screening data if they were obtained within 90 days from the date of re-consent.

If determined to be eligible, the subject will be enrolled.

The following observations and examinations will be performed.

(1) Appearance (also take photographs of the lesion site)
(2) Laboratory tests (red blood cell count, hemoglobin, hematocrit, white blood cell count, platelet count, white blood cell fraction [neutrophils, lymphocytes, eosinophils, basophils, monocytes], PT, APTT, PT-INR, fibrinogen, antithrombin III, FDP, D-dimer, Na, K, Cl, Ca, IP, BUN, Cre, UA, AST, ALT, LDH, γ-GTP, T.Bil, TP, Alb, ALP, CK, TG, T-chol, HDL-chol, AMY, CRP, Glu)
(3) Urinalysis (urine qualitative: sugar, protein, urobilinogen, occult blood, white blood cells, pH, specific gravity)
(4) Subjective symptoms (pain)
(5) MRI (CT and ultrasound examination, if necessary)
(6) QOL (including ADL)
(7) Adverse event
(8) Concomitant medications

6.2.2　1 day before administration

The following observations and examinations will be performed on the day before the sclerotherapy (administration of the study drug). The subject should be hospitalized.

(1) Vital signs (blood pressure, pulse rate, body temperature, SpO_2_ )
(2) Body weight
(3) Subjective symptoms (pain)
(4) Adverse event
(5) Concomitant medications

6.2.3　Treatment period

6.2.3.1　Date of administration

The investigational drug will be administered within 1 month (within 30 days) after obtaining consent from the subject or the surrogate.
 If the drug is not administered after one month from obtaining consent, re-consent will be obtained to confirm the subject's or surrogate's willingness. The subject should be hospitalized.

The following observations and examinations will be performed.

(1) Administration of investigational drug

At each site, the investigational drug is received from the investigational drug manager.

Based on body weight on the day prior to administration, administer the investigational drug at a maximum dose of 0.4 mL/kg of 5% monoethanolamine oleate (extreme dose is 30 mL). The dose (mL) administered should also be recorded.

(2) Adverse events

(3) Concomitant medications

6.2.3.2　Day after administration (Tolerance ±0)

The subject should be hospitalized and will undergo the following observations and examinations will be performed.

(1) Vital signs (blood pressure, pulse rate, body temperature, SpO_2_ )
(2) Laboratory tests (red blood cell count, hemoglobin, hematocrit, white blood cell count, platelet count, white blood cell fraction [neutrophils, lymphocytes, eosinophils, basophils, monocytes], PT, APTT, PT-INR, fibrinogen, antithrombin III, FDP, D-dimer, Na, K, Cl, Ca, IP, BUN, Cre, UA, AST, ALT, LDH, γ-GTP, T.Bil, TP, Alb, ALP, CK, TG, T-chol, HDL-chol, AMY, CRP, Glu)
(3) Urinalysis (urine qualitative: sugar, protein, urobilinogen, occult blood, white blood cells, pH, specific gravity)
(4) Subjective symptoms (pain)
(5) Adverse event
(6) Concomitant medications

6.2.3.3　3 days after administration (Tolerance ±1 day)

If there are no adverse events (abnormal liver function, abnormal renal function) associated with the administration of the investigational drug, the patient will be discharged from the hospital in principle, and the following observations and examinations will be performed prior to discharge. The allowable range of observation days is ±1 day.

(1) Vital signs (blood pressure, pulse rate, body temperature, SpO_2_ )
(2) Subjective symptoms (pain)
(3) Adverse event
(4) Concomitant medications

6.2.3.4 　2 weeks after administration (Tolerance ±5 days)

The following observations and examinations will be performed. The allowable range of observation days is ±5 days.

The subject will be treated as outpatients.

(1) Vital signs (blood pressure, pulse rate, body temperature, SpO_2_ )
(2) Subjective symptoms (pain)
(3) Adverse event
(4) Concomitant medications

6.2.3.5．　4 weeks after administration (Tolerance ±5 days)

The following observations and examinations will be performed. The allowable range of observation days is ±5 days.

The subject will be treated as outpatients.

(1) Vital signs (blood pressure, pulse rate, body temperature, SpO_2_ )
(2) Subjective symptoms (pain)
(3) QOL (including ADL)
(4) Adverse event
(5) Concomitant medications

6.2.3.6　3 months after administration (12 weeks after administration) (Tolerance ±7 days)

The following observations and examinations will be performed. The allowable range of observation days is ±7 days. However, for MRI imaging only, the allowable range is ±14 days.

The subject will be treated as outpatients.

(1) Vital signs (blood pressure, pulse rate, body temperature, SpO_2_ )
(2) Appearance (also take photographs of the lesion site)
(3) Subjective symptoms (pain)
(4) MRI
(5) QOL (including ADL)
(6) Adverse event
(7) Concomitant medications

6.2.3.7　(In case of additional treatment) Date of additional administration - 3 months after additional administration (12 weeks after additional administration)

If additional treatment is administered, observation and examination should be performed according to the above "6.2.3.1 Date of administration" through "6.2.3.6 Three months after administration (12 weeks after administration)" starting from the date of the additional administration.

6.2.4　Discontinuation

6.2.4.1　Time of discontinuation

If during the treatment/observation period of a subject (from the date of administration to 3 months of administration), any of the criteria in "7.1 Termination (discontinuation) of treatment of individual subjects" or "4.1.12 Criteria for discontinuation of investigational drug " are met, the administration of the subject will be discontinued (if the investigational drug is being administered) and the following observations and examinations will be performed to ensure the safety of the subject.

The subject shall be hospitalized or outpatients, depending on the situation.

(1) Vital signs (blood pressure, pulse rate, body temperature, SpO_2_ )
(2) Appearance (also take photographs of the lesion site)
(3) Laboratory tests (red blood cell count, hemoglobin, hematocrit, white blood cell count, platelet count, white blood cell fraction [neutrophils, lymphocytes, eosinophils, basophils, monocytes], PT, APTT, PT-INR, fibrinogen, antithrombin III, FDP, D-dimer, Na, K, Cl, Ca, IP, BUN, Cre, UA, AST, ALT, LDH, γ-GTP, T.Bil, TP, Alb, ALP, CK, TG, T-chol, HDL-chol, AMY, CRP, Glu)
(4) Urinalysis (urine qualitative: sugar, protein, urobilinogen, occult blood, white blood cells, pH, specific gravity)
(5) Subjective symptoms (pain)
(6) QOL (including ADL)
(7) Adverse event
(8) Concomitant medications

6.2.4.2　3 months after administration (12 weeks after administration) <discontinued cases> (Tolerance ±7 days)

The following observations and examinations will be performed. The allowable range of observation days is ±7 days. However, for MRI imaging only, the allowable range is ±14 days.

The subject will be treated as outpatients.

(1) Vital signs (blood pressure, pulse rate, body temperature, SpO_2_ )
(2) Appearance (also take photographs of the lesion site)
(3) Subjective symptoms (pain)
(4) MRI
(5) QOL (including ADL)
(6) Adverse event
(7) Concomitant medications

6.2.4.3　(In case of additional treatment) 3 months after additional administration (12 weeks after additional administration) <Discontinued cases>

If administration is discontinued after additional treatment, observation and examination will be performed according to the above "6.2.4.1 Time of discontinuation " and "6.2.4.2 3 months after administration (12 weeks after administration) <discontinued cases> " starting from the date of additional administration.

7.　Termination of the subject's treatment or evaluation

7.1　Termination (discontinuation) of treatment of individual subjects

The investigator and subinvestigator will discontinue the administration of the investigational drug and discontinue this clinical trial for subjects who fall into any of the following categories. If an adverse event has occurred at the time of discontinuation, the investigator and subinvestigator will take appropriate measures for the adverse event and follow up the outcome of the adverse event to the extent possible, with a deadline of the end date of this entire clinical trial (scheduled last observation date for all subjects).

- The subject or surrogate withdraws consent for the administration of the investigational drug to the subject or for participation in the clinical trial (for any reason)
- If continued participation in the clinical trial is likely to endanger the subject
- When "4.1.12 Criteria for discontinuation of investigational drug " is applicable.
- If a subject is found not to meet eligibility criteria
- If the investigator or subinvestigator determines that the subject or surrogate is unable to comply with the study protocol
- If the subject is unable to come to the hospital due to reasons on the subject's side (e.g., relocation, hospital transfer, etc.)
- If the subject becomes pregnant
- Other medical reasons (e.g., surgery, adverse events, other diseases, etc.) that make it impossible to administer the investigational drug
- When the investigator or subinvestigator determines that it is appropriate to discontinue the clinical trial for other reasons

7.2　Discontinuation of the clinical trial

If any of the following reasons apply, the Coordinating Committee shall, from time to time, suspend the entire clinical trial or the clinical trial at a specific site. If necessary, the Efficacy and Safety Evaluation Committee will be consulted, and a decision on whether or not to continue the clinical trial (continuation, suspension, or discontinuation) will be made based on the recommendation of the Efficacy and Safety Evaluation Committee. Details of the Efficacy and Safety Evaluation Committee are separately stipulated in the "Procedures for the Efficacy and Safety Evaluation Committee.

- Efficacy or safety information that may have a significant impact on the continuation of the clinical trial comes to light
- When there is a violation of GCP or a violation of the study protocol that hinders the proper conduct of the clinical trial
- Serious or continuing noncompliance by the investigator or subinvestigator is discovered
- When it becomes impossible to continue the clinical trial due to a change in the principal investigator
- When it is deemed impossible to reach the planned target number of cases (discontinuation of the entire clinical trial or at a specific site)
- When the site no longer meets the requirements to properly conduct the clinical trial
- If the IRB decides to discontinue the clinical trial
- In the event of a recommendation from the regulatory authority to cease and desist
- When the Coordinating Committee determines that it is appropriate to discontinue the clinical trial for other reasons

8. Statistical analysis

Due to differences in disease characteristics, cystic and diffuse lesions will be analyzed separately as separate cohorts for all endpoints.

8.1　Analysis population

8.1.1　Definition of analysis population

The population to be analyzed is defined as follows.

(1) Full analysis set (FAS): all subjects who received the study drug

(2) Per protocol set (PPS): The subjects in the FAS, excluding those with the following serious violations of the study protocol regulations in terms of study methods, concomitant therapy, etc. The PPS will be set for the purpose of confirming the stability of the primary analysis for the efficacy endpoints.

・Inclusion criteria violation

・Exclusion criteria violation

・Significant deviations from the study protocol that could affect the evaluation of efficacy

(3) Safety analysis set (SAS): Same as FAS

8.1.2　Endpoints and analysis populations

The analysis population for each endpoint is shown in Table 8.1.2-1. The primary analysis population for the efficacy endpoints is the FAS.

Table 8.1.2-1 Evaluation items and analysis population

| Evaluation item | Analysis population | |
| --- | --- | --- |
|  | Cystic lesion | Diffuse lesion |
| Subject basic information | FAS | FAS |
| Primary endpoint | FAS, PPS | FAS, PPS |
| Secondary endpoints | FAS, PPS | FAS, PPS |
| Safety endpoints | SAS | SAS |

8.1.3　Handling of individual cases

Prior to data fixed, the case review meeting is held, and the Coordinating Committee and the statistical analysis manager will discuss and decide on the handling of individual subjects.

8.2　Analysis policy

8.2.1　General policy

The statistical analysis manager shall prepare a statistical analysis plan prior to data fixed, and shall perform all analyses in accordance with the statistical analysis plan.

8.2.2　Efficacy analysis

8.2.2.1　Analysis for primary endpoint

For the number of subjects who achieve at least 20% reduction from baseline (before administration) in the volume of venous malformations at 3 months of treatment with the investigational drug, if the null hypothesis is "the proportion of the population achieving at least 20% reduction from baseline in the volume of venous malformations at 3 months of treatment with the investigational drug is less than 20%", a statistical test based on a binomial distribution are performed. The significance level will be 2.5% one-sided. The 95% confidence interval between the proportion of subjects who achieve at least 20% reduction from baseline in the volume of venous malformations at 3 months of treatment with the investigational drug and the proportion based on the score test will also be estimated.

8.2.2.2　Analysis for secondary endpoints

8.2.2.2.1　Analysis for important secondary endpoints

Estimate the median -1-fold change from baseline (pre-dose) to 3 months after sclerotherapy and its 95% confidence interval for the symptom (pain) score associated with the target lesion at 3 months after sclerotherapy. Also, if the null hypothesis is "the median -1-fold change from baseline to 3 months post-sclerotherapy in the population is less than or equal to 0." and a Wilcoxon signed-rank test is performed. The significance level is set at 2.5% one-sided.

8.2.2.2.2　Analysis for other secondary endpoints

(1) Estimate the median and 95% confidence interval for the reduction rate (volume) of the target lesion from baseline (before administration) at 3 months after sclerotherapy. Also, if the null hypothesis is "The median reduction rate (volume) of the target lesion at 3 months after sclerotherapy is less than or equal to zero.", the Wilcoxon signed-rank test is performed. The significance level is set at 2.5% one-sided.

(2) Estimate the median -1-fold change from baseline (before administration) to one month after sclerotherapy and its 95% confidence interval for the symptom (pain) score associated with the target lesion at one month (4 weeks) after sclerotherapy. Also, if the null hypothesis is "the median -1-fold change from baseline to one month after sclerotherapy in the population is less than or equal to zero.", the Wilcoxon signed-rank test is performed. The significance level is set at 2.5% one-sided.

(3) Estimate the median change from baseline (before administration) to 3 months after sclerotherapy and its 95% confidence interval for the QOL score. Also, if the null hypothesis is "the median change from baseline to 3 months after sclerotherapy in the population is less than or equal to zero.", the Wilcoxon signed-rank test is performed. The significance level is set at 2.5% one-sided.

8.2.2.3　Analysis for other endpoints

Estimate the percentage of each category (much improvement, improvement, unchanged, and worsening) for change in appearance at 3 months after sclerotherapy. We will also estimate the percentage of much improvement or improvement and its 95% confidence interval, with the null hypothesis being "the percentage of cases with much improvement or improvement is less than 20%." and then perform a test based on the binomial distribution. The significance level is set at 2.5% one-sided.

8.2.2.4　Subgroup analysis

For "8.2.2.1 Analysis for primary endpoint" and "8.2.2.2 Analysis for secondary endpoints," a subpopulation analysis will be conducted for four age categories: 5 years and younger, 6 to 11 years, 12 to 14 years, and 15 years and older.

8.2.3　Safety analysis

(1) For each adverse event and adverse drug reaction, the number of subjects and the incidence rate are shown. The number of subjects and the incidence rate by organ, symptoms and findings, and grade should be tabulated. For SAEs, the number of subjects and the incidence rate should be indicated and listed.

(2) Summarize the summary statistics of laboratory measurements by observational period.

8.3　Handling of missing values

No inputation will be performed in case of missing data.

8.4　Interim analysis

No interim analysis is planned.

8.5　Subject background

For basic information on subjects, for each measure, describe the summary statistics for each lesion (cystic or diffuse). For continuous scales, the number of cases, mean, standard deviation, minimum, median, and maximum values are calculated. For nominal and ordinal scales, category frequencies and proportions are presented.

8.6　Target number of cases and rationale for setting

8.6.1　Target number of cases

44 cases (22 cystic lesions, 22 diffuse lesions)

8.6.2　Basis for setting

8.6.2.1 　Cystic lesions

For the difficult-to-resect venous malformations (cystic lesions) that are the subject of this trial, no improvement is expected as a natural history. However, we set the threshold for this study at 20% because we believe that the percentage of subjects who achieve CR (target lesion disappearance) and PR (target lesion volume reduction of at least 20%), which were defined as the achievement criteria, must improve by at least 20% when the treatment is administered.

Studies evaluating MRI-based monoethanolamine oleate for difficult-to-resect venous malformations are limited. Kaji et al.^10)^ reported a study of lesion reduction rates using MRI in subjects with venous malformations included. From the box-and-whisker diagram shown for venous malformations (60 cases), it can be read that the median is about 25%. Since the median value corresponds to the 50th percentile of the distribution, the percentage of subjects with a reduction rate of 20% or greater would be expected to be greater than 50% as read from the figure. The study by Alexander et al.^13)^ of the reduction rate using MRI for venous malformations also suggests that the proportion of subjects with a reduction rate of 20% or greater would be expected to be 50% or greater. Kaji et al. converted lesion reduction rate from area (Alexander et al. converted from volume), which is not the same as the evaluation of target lesion volume set for this trial, but we considered the reduction rates in these studies to be appropriate for this trial and expected the proportion of CR and PR to be 50% or more.

In this setting, the number of cases needed to obtain 80% power in a test based on a binomial distribution with a significance level of 2.5% one-sided is 19. Considering dropout cases, the target number of cystic lesion patients was set at 22 cases. In order to ensure a certain number of cases in the younger age group in actual clinical practice, the target number of cases was set for each age group as follows
 (under 15 years old: target 10 cases).

8.6.2.1 　Diffuse lesions

Patients with diffuse lesions are thought to be more numerous than those with cystic lesions, but considering the feasibility of the study, taking into account the variation in the number of patients from year to year, we considered it appropriate to have about the same number of patients as the target number of patients with cystic lesions, which is 22 cases including dropouts (under 15 years old: target 10 cases).

8.7　Trial period

Enrollment period: January 1, 2021 - March 31, 2023

Trial period: January 1, 2021 - June 30, 2023

9.　Informed consent, data protection, and clinical trial records

9.1　Ethics and protection of subjects

This clinical trial will be conducted in compliance with the protocol, the ethical principles of the Declaration of Helsinki, and in accordance with GCP (Ministry of Health and Welfare Ordinance No. 28, dated March 27, 1997) and related regulations.

The responsibility for standard medical care (preventive, diagnostic, and therapeutic actions) rests with the treating physician.

The investigators and subinvestigators must also promptly report to the Coordinating Committee any emergency safety measures taken to protect subjects from imminent danger and all serious violations of the study protocol and GCP.

9.2　Approval of the clinical trial, provision of information to the subject or surrogate, and informed consent

Only after all necessary legal documents have been reviewed and approved by the respective IRBs, this clinical trial will begin. The same situation applies to amendments to the protocol.

Written consent must be obtained from each subject or surrogate prior to participation in this study in accordance with GCP and regulatory and statutory requirements. The consent form must be signed and dated by each person, and the consent form and consent explanatory document must be kept by the investigator and subinvestigator as part of the records of this study. In addition, a copy of the signed consent form and the consent explanatory document must be delivered to the subject or surrogate.

The investigators and subinvestigators must provide adequate explanations to the subject or the surrogate, using a consent document prepared using language and expressions that can be understood by non-specialists to the greatest extent possible. They must also give the subject or surrogate a sufficient amount of time to consider participation in the study, confirm that the subject or surrogate understands the content, and obtain free will consent in writing from the subject or surrogate using the consent form. The investigator and subinvestigator must sign or write their names/stamps on the consent form and date it respectively. If a clinical research coordinator provides a supplementary explanation, the clinical research coordinator must also sign or write his/her name/stamp on the form and date it respectively.

If new significant findings are obtained, re-consent must be obtained.

9.3　Data quality assurance

The IRB or regulatory agency may conduct a quality assurance audit/inspection of this clinical trial. The auditor will have access to all medical records related to this clinical trial, investigator and subinvestigator’s materials and correspondence related to the clinical trial, and subject or surrogate consent forms.

9.4　Record

Individual subject data will be recorded in the CRF. Refer to "4.1.14 Investigational drug management" for the management of investigational drugs.

9.4.1　Source documents

Source documents (source data) for this clinical trial shall be the following documents and materials. Data stored in electronic medical records shall also be considered source documents.

・Records of consent of the subject or surrogate

・Records of information provided to the subject or surrogate

・Medical records, etc. (medical records, nursing records, laboratory data, prescription records, etc.)

・Records of imaging examination results (e.g., imaging examination films)

・Records related to the administration of investigational drug

Source documents will be kept at each site.

The data recorded in the CRF must be consistent with the source documents, and if they are not consistent with the source documents, an appropriate explanation must be provided. For some clinical trials, investigators and subinvestigators may need to obtain medical records prior to the start of the trial, transfer records, etc. The most recent medical records at the time of the trial must be available for inspection. All data recorded in the CRF must be based on the source documents.

9.4.2　Direct access to source documents

The investigator and subinvestigator/sites shall be open to monitoring and audits, IRB investigations and regulatory inspections, and shall have direct access to all source documents related to the clinical trial. In addition, the CRF and all source documents (including copies of progress notes, laboratory and other test results) must be available at all times for monitoring, investigator review and regulatory inspection. Audit personnel may examine all CRFs and written consent statements. The accuracy of the data should be checked against the source documents as described in "9.4.1 Source documents".

9.4.3　Retention period for records

The site must retain the source documents and essential documents for the period specified in the GCP.

9.5　Maintenance of secrecy

Each subject's medical information obtained in this clinical trial will be confidential and will not be disclosed to any third party, with the following exceptions. The confidentiality of each subject will be ensured by the use of a subject identification number.

Investigators and subinvestigators, as well as other health care providers, may provide clinical trial data. They should also be available for inspection by the Coordinating Committee, the IRB and regulatory authorities.

9.6　Termination of clinical trials

Between the date of the last observation on the entire subject population and the completion of the Clinical Study Report (CSR), the completion of work associated with the clinical trial at each site is defined as the end of the clinical trial at that site.

When a clinical trial is terminated, the investigator shall notify the head of the site in writing, and the head of the site shall promptly notify the IRB of the termination in writing.

9.7　Deviations from the study protocol

The investigators and subinvestigators must record all deviations from the study protocol for any reason. If the investigator deviates from or changes the protocol in order to avoid immediate risk to the subject or for other compelling medical reasons, the investigator shall report the details of and reasons for the deviation or change to the IRB of the site as soon as possible and obtain its approval.

10.　Protocol amendment

When the investigator and the Coordinating Committee become aware of important information for the proper conduct of a clinical trial, such as matters related to the quality, efficacy and safety of the investigational drug, they shall consult with the other investigators, revise the protocol as necessary, obtain agreement among the investigators, and have it reviewed by the IRB at each site. If necessary, the Efficacy and Safety Evaluation Committee shall be consulted, and a decision on whether or not to revise the protocol shall be made based on the recommendation of the Efficacy and Safety Evaluation Committee. Details of the Efficacy and Safety Evaluation Committee are separately stipulated in the "Procedures for the Efficacy and Safety Evaluation Committee.

If it is necessary to revise the consent forms and explanation documents in accordance with the revision of the protocol, the investigator shall promptly revise the consent forms and explanation documents, report the revised contents to the head of the site, and submit the revised consent forms and explanation documents to the IRB for review. If the consent forms and explanation documents are revised, the investigator and the subinvestigator shall inform the subject or the surrogate of the revised contents, confirm the subject's willingness to continue participating in the clinical trial, and obtain consent again using the revised consent forms and explanation documents.

11.　Funds and conflicts of interest

11.1　Funds (money)

This clinical trial will be funded by the Japan Agency for Medical Research and Development (AMED).

In addition, investigational drug and safety information (foreign case reports [unknown/serious adverse reactions], action reports, research reports, etc.) will be provided by the investigational drug provider, Fuji Chemical Industry Co., Ltd.

11.2　Conflict of interest

The possibility of conflicts of interest between the Coordinating Committee, investigators and subinvestigators, and the investigational drug provider, Fuji Chemical Industry Co., Ltd. regarding the conduct and publication of this clinical trial shall be reviewed by the conflict of interest committee of each site to maintain fairness regarding the conflicts of interest in this clinical trial.

Fuji Chemical Industry Co., Ltd. will provide the investigational drug and safety information of it, but will not be involved in the collection, management, or analysis of data in this clinical trial, and will not be in a situation where this will affect the results of the study.

12.　Payment of money, etc. and insurance

12.1　Payment of money, etc.

Since the investigational drug is provided free of charge by the pharmaceutical company, there is no cost burden on the subject. The costs related to medical treatment during the clinical trial period, other than the cost of the investigational drug, will be paid by the subject.

In addition, a reimbursement (compensation for cooperating in the clinical trial) shall be paid to the subject or surrogate for each inpatient admission and each outpatient visit. This payment will be made through each site in accordance with their rules.

12.2　Health damage compensation

If a subject suffers health problems as a result of participating in this clinical trial, the site will take necessary and appropriate measures such as providing a medical treatment. If the adverse health effects are judged to be due to the intentional or gross negligence of the subject or the surrogate, compensation may not be provided.

The investigator-initiated clinical trial insurance will be purchased as a measure to cover any liability for compensation for health damage resulting from this clinical trial. The President of Kyorin University shall be the policyholder, and the investigators, subinvestigators and site shall be insured by the insurance. The investigators and subinvestigators shall also be covered by the physician liability insurance.

13.　Publication arrangements

The results of this clinical trial, regardless of the outcome, will be presented at (1) an academic conference and (2) submitted to a medical journal as the outcome of a collaborative study. In such cases, the results shall be co-authored by a member of the Coordinating Committee, the investigator(s) at the site of the study, and the person(s) responsible for statistical analysis. However, in principle, the results of the clinical trial must not be published before the completion of the CSR.

In addition, the results of subjects enrolled in the trial shall not be published individually prior to publication as a collaborative study.

14.　Organization

The organization of this trial is described in Appendix 1.

15.　References

1 Kang GB, et al. The usefulness of surgical treatment in slow-flow vascular malformation patients Arch Plast Surg. 2017;44:301-7.
2 Jo JY, Chin J, Park PH, et al: Cardiovascular collapse due to right heart failure following ethanol sclerotherapy. Korean J Anesthesiol. 2014;66:388-91 .
3 Shimochi H, Hidaka K, Yanagawa S, et al: A case of a child with two cardiac arrests during vascular sclerotherapy with polidocanol. Anesthesia. 2005;54:57-9.
4 Horbach SER, et al. Sclerotherapy for low-flow vascular malformations of the head and neck: a systematic review of sclerosing agents J Plast Reconstr. Aesthet Surg. 2016;69:295-304.
5 Ozaki M, et al. Efficacy and evaluation of the safety of sclerosants for intramuscular venous malformations: clinical and experimental studies. Scand J Plast Reconstr Surg Hand Surg. 2010;44:75-87.
6 Hoque S, et al: Treatment of venous malformations with ethanolamine oleate: a descriptive study of 83 cases. Pediatr Surg Int. 2011;27:527-31.
7 Mulliken JB, Oxford university Press. 2013
8 Choi YH, et al: Craniofacial cavernous venous malformations: percutaneous sclerotherapy with use of ethanolamine oleate. J Vasc Interv Radiol. 2002; 13:475-82. J Vasc Interv Radiol. 2002; 13:475-82.
9 T. Nomura, et al: Our treatment strategy for hemodynamic venous malformations: indications and limitations of sclerotherapy. Phlebology. 2008;19:161-8.
10 Kaji N, et al: Experience of sclerotherapy and embolosclerotherapy using ethanolamine oleate for vascular malformations of the head and neck. Scand J Plast Reconstr Surg Hand Surg. 2009;43:126-36.
11 Costa JR, et al: Sclerotherapy for vascular malformations in the oral and maxillofacial region: treatment and follow-up of 66 lesions. J Oral Maxillofac Surg. 2011;69:e88-e92.
12 Hoque S, et al: Treatment of venous malformations with ethanolamine oleate: a descriptive study of 83 cases. Pediatr Surg Int. 2011;27:527-31.
13 Alexander MD, et al. Percutaneous sclerotherapy with ethanolamine oleate for venous malformations of the head and neck. NeuroIntervent Surg 2014;6: 695-8. 695-8.

Revision History

| version number | Creation/Revision Date | Reason for Revision/Contents |
| --- | --- | --- |
| Version 1.0 | June 24, 2020 | newly enacted |
| Version 1.1 | July 17, 2020 | Clarification of the timing and number of additional doses, deletion of some secondary evaluation items, improvement of abbreviations, improvement of wording, unification of terminology, and correction of typographical errors. |
| Version 1.2 | September 14, 2020 | Clarification of rating scale by patient age group, addition of rating scale for guardians, improvement of content and timing of observation, improvement of descriptions, correction of typos and errors. |
| Version 1.3 | January 6, 2021 | Clarification of the following  The criteria for selection, the method of evaluation at the time of additional treatment, the rationale for safety information (background information), the duration of concomitant medications/adjunctive therapy, the period during which concomitant medications are prohibited, the criteria for adoption of preadministration data, the timing of weight measurements, and the number of months."  Addition of inspection items, addition of notes, reordering of references, word maintenance, word correction, terminology unification, and correction of errors |
| Version 1.4 | April 1, 2021 | Specify the QOL evaluation, add a part of the investigation period for QOL evaluation, change the anesthesia method, clarify patients eligible for additional treatment, clarify the definition of abnormal values to be reexamined in laboratory tests, etc., change the name of the institution, delete job titles, change the wording, etc. |
| Edition 1.5 | June 18, 2021 | Change of anesthesia method for children, clarification of provisions for children and adults in this clinical trial |
| Version 1.6 | October 21, 2021 | Addition of handling of MRI images at the time of obtaining re-consent, and change of conditions for timing of MRI imaging. |
| Version 1.7 | October 26, 2022 | Change of tables according to the type of questionnaire, maintenance of descriptions, addition of abbreviations, correction of errors |
| Version 1.8 | December 7, 2022 | Extension of the expected period of subject enrollment |
